# Genome-wide interaction study implicates *VGLL2* and alcohol exposure and *PRL* and smoking in orofacial cleft risk

**DOI:** 10.1101/2020.10.26.20219980

**Authors:** Jenna C. Carlson, John R. Shaffer, Fred Deleyiannis, Jacqueline T. Hecht, George L. Wehby, Kaare Christensen, Eleanor Feingold, Seth M. Weinberg, Mary L. Marazita, Elizabeth J. Leslie

## Abstract

Non-syndromic cleft lip with or without cleft palate (NSCL/P) is a common birth defect, affecting approximately 1 in 700 births. NSCL/P has complex etiology including several known genes and environmental factors; however, known genetic risk variants only account for a small fraction of the heritability of NSCL/P. It is commonly suggested that gene-by-environment (G×E) interactions may help explain some of the “missing” heritability of NSCL/P. We conducted a genome-wide G×E interaction study in European cases and controls with three common maternal exposures during pregnancy: alcohol, smoking, and vitamin use using a two-stage design. After selecting 127 loci with suggestive 2df tests for gene and G x E effects, 40 loci showed significant G x E effects after correcting for multiple tests. Notable interactions included SNPs of 6q22 near VGLL2 with alcohol and 6p22.3 near PRL with smoking. These interactions could provide new insights into the etiology of CL/P and new opportunities to modify risk through behavioral changes.

## INTRODUCTION

Interest in identifying the causal factors for birth defects, including orofacial clefts (OFCs) can be traced back centuries (1) and has often involved a debate as to the contribution of genetic versus environmental risk factors(2). Evidence for a multifactorial model for OFC etiology originated with Fraser’s early studies exposing pregnant mice to cortisone showing that the incidence of corticosteroid induced cleft palate varied by strain. The multifactorial model is favored in human nonsyndromic OFCs, where twin and family studies provide evidence for a strong, but incomplete, genetic component. Many environmental factors have been postulated to modify risk of OFCs including maternal medications, smoking or alcohol consumption(3, 4), nutrition (5), obesity (6), gestational diabetes (7), and occupational exposures. Of these, cigarette smoking, alcohol consumption, and folic acid supplementation are the most widely studied, but, except for smoking(8), these exposures have inconsistent results in epidemiological studies. Maternal smoking, on the other hand, has been shown to increase risk consistently across many studies with similar effect sizes(9).

Over the last ten years, 20 independent genome-wide association studies (GWAS) or meta-analyses have identified at least 50 loci associated with OFCs (10). Cumulatively, these loci are estimated to account for 25-30% of the heritable risk attributed to additive genetic effects. Other approaches are needed to identify the “missing” heritability, some of which may be attributed to interaction effects. Leveraging published data from GWAS studies, gene-environment interaction (GxE) studies have become a popular approach to further elucidate OFC risk.

Following one such GWAS study in European and Asian case-parent trios (Beaty 2010), candidate gene and pathway-based GxE analyses have suggested interactions between smoking and *RUNX2* (11), and between multiple exposures and *BMP4* (12). Genome-wide GxE using a variety of statistical approaches have identified interactions for maternal smoking, alcohol, or folate supplementation in cleft palate (13-15) and cleft lip with or without cleft palate (16, 17).

In this study, we performed genome-wide GxE analyses in a European case-control sample from the Pittsburgh Orofacial Cleft Study to identify interactions between genetic variants and maternal smoking, alcohol consumption, or vitamin supplementation during the periconceptional period that influence risk of OFCs.

## MATERIALS AND METHODS

### Study Samples and Genotyping

The sample for these analyses was derived from a larger multiethnic OFC cohort, which has been previously described (18). Briefly, participants were recruited from 18 sites worldwide as part of ongoing genetic studies conducted by the University of Iowa and the University of Pittsburgh Center for Craniofacial and Dental Genetics. All sites obtained Institutional Review Board approval both locally and at the University of Iowa or the University of Pittsburgh. All participants gave informed consent. These data are available through dbGaP (accession number: phs000774.v2.p1).

This large multiethnic cohort contains OFC-affected probands and their unaffected relatives in addition to controls without a family history of OFC or other craniofacial anomalies. Additionally, data were obtained on three maternal periconceptional exposures: alcohol use, smoking, and vitamin use. Periconceptional exposures were gathered from maternal self-report of alcohol use, personal smoking, or vitamin use during the three months prior to pregnancy and during the first trimester for each child.

From this larger cohort, a subset of unrelated cases and controls of European ancestry and with complete data for the three environmental exposure measures was drawn. It consisted of 344 NSCL/P cases and 194 controls of European descent from Denmark, Hungary, and the United States (Table 1).

**Table 1.** Characteristics of Sample. Sample size (column percent) comparing environmental exposure status between cases and controls within each recruitment site.

The genotyping, quality control procedures, imputation, and generation of principal components of ancestry (PCAs) have been previously described (18). Briefly, samples were genotyped using the Illumina HumanCore + Exome chip (Illumina Inc., San Diego, CA), with approximately 97% of the genotyped SNPs passing quality control filters, resulting in a total of 539,473 genotyped SNPs. Genotype imputation for an additional 34,985,077 unobserved polymorphisms was performed using IMPUTE2 software with the multiethnic 1,000 Genomes Project Phase 3 reference panel (19). Imputed genotype probabilities were converted to most-likely genotypes using GTOOL; only most-likely genotypes with probabilities >0.9 were retained for statistical analysis. Imputed SNPs with INFO scores <0.5 or those deviating from Hardy-Weinberg equilibrium (p<1×10^−4^ in a set of unrelated European controls) were also excluded from the analysis. The University of Washington, Genetics Coordinating Center released a full online report on the data cleaning, quality assurance, ancestry analyses, and imputation for this study (http://www.ccdg.pitt.edu/docs/Marazita_ofc_QC_report_feb2015.pdf, last accessed April 25, 2016). Additionally, we excluded variants with a minor allele frequency < 10%, as the power to detect interaction effects among low-frequency variants is very limited and analysis of such variants is prone to type 1 errors. After these filtering steps, 5,165,675 were included in the genome-wide interaction analyses. Population structure was measured through generation of principal components of ancestry using the R package SNPRelate following the approach introduced by Patterson, Price, and Reich (20).

### Statistical Analyses

To assess the association between genetic variants and NSCL/P that may or may not be modified by an environmental factor, we used the strategy laid out in Kraft et al 2007 for each of the three environmental factors (21). First, a 2 degree of freedon (2df) joint test of the gene (G) and gene-environment interaction (GE) effects was employed genome-wide using logistic regression, adjusting for five principal components of ancestry. This provides a sensitive test for two scenarios simultaneously: (1) where there is a genetic effect that is similar across all environmental strata, and (2) where there is a genetic effect that is specific to a specific environmental stratum. Then, the GE effect was interrogated for any variants demonstrating at least suggestive results from the G-GE joint test (i.e., p<1×10^−4^) to identify regions in which there is a genetic effect differing across environmental stratum. We used a Bonferroni-adjusted significance threshold for examining the GE effect, adjusting for the number of regions with suggestive results from the G-GE joint test. All analyses were conducted in PLINK v1.9 (22).

## RESULTS

As a follow-up study to previous GWAS identifying marginal gene effects (G) increasing risk for NSCL/P, this study focused on identifying G x E interactions. We carried out a two-stage analysis, where we first examined the 2df joint test of G and G x E (GE). We observed suggestive evidence of association in the joint G-GE test for 127 loci. Several loci had multiple SNPs with p<1×10^−4^ in each of the three analyses. These included 8q24, one of the most reliably associated loci with NSCL/P in European populations (10). As expected, this locus showed strong G effects, but did not have a significant GE effect (p>0.05). A locus on 2p23 with similarly strong G effects but no GE effects was identified in all three analyses but to our knowledge has not been reported before in GWAS of NSCL/P.

To identify gene-environment interactions, we next examined the GE effect alone for each variant with p<1×10^−4^ in the joint test. 40 regions (212 SNPs) had a GE p-value less than 3.9×10^−4^ (i.e., Bonferroni correction for 127 loci) (Table 2), although none had a genome-wide significant GE p-value. Among these regions, we observed a statistically significant interaction effect between rs706954, a variant <5kb downstream of *VGLL2*, and periconceptional alcohol use (p_GE_=4.62×10^−6^). The minor allele (G) of rs706945 was associated with higher odds of NSCL/P within individuals with periconceptional alcohol use, but lower odds with individuals without periconceptional alcohol use (Figure 2).

**Table 2.** Results for top-associated variants. Columns: CHR, BP, SNP, A2 (major), A1 (minor), MAF, imputed, P.2df, P.gxe, OR (G when E=0), OR (G when E=1), scan, cytoband

**Figure 1.**
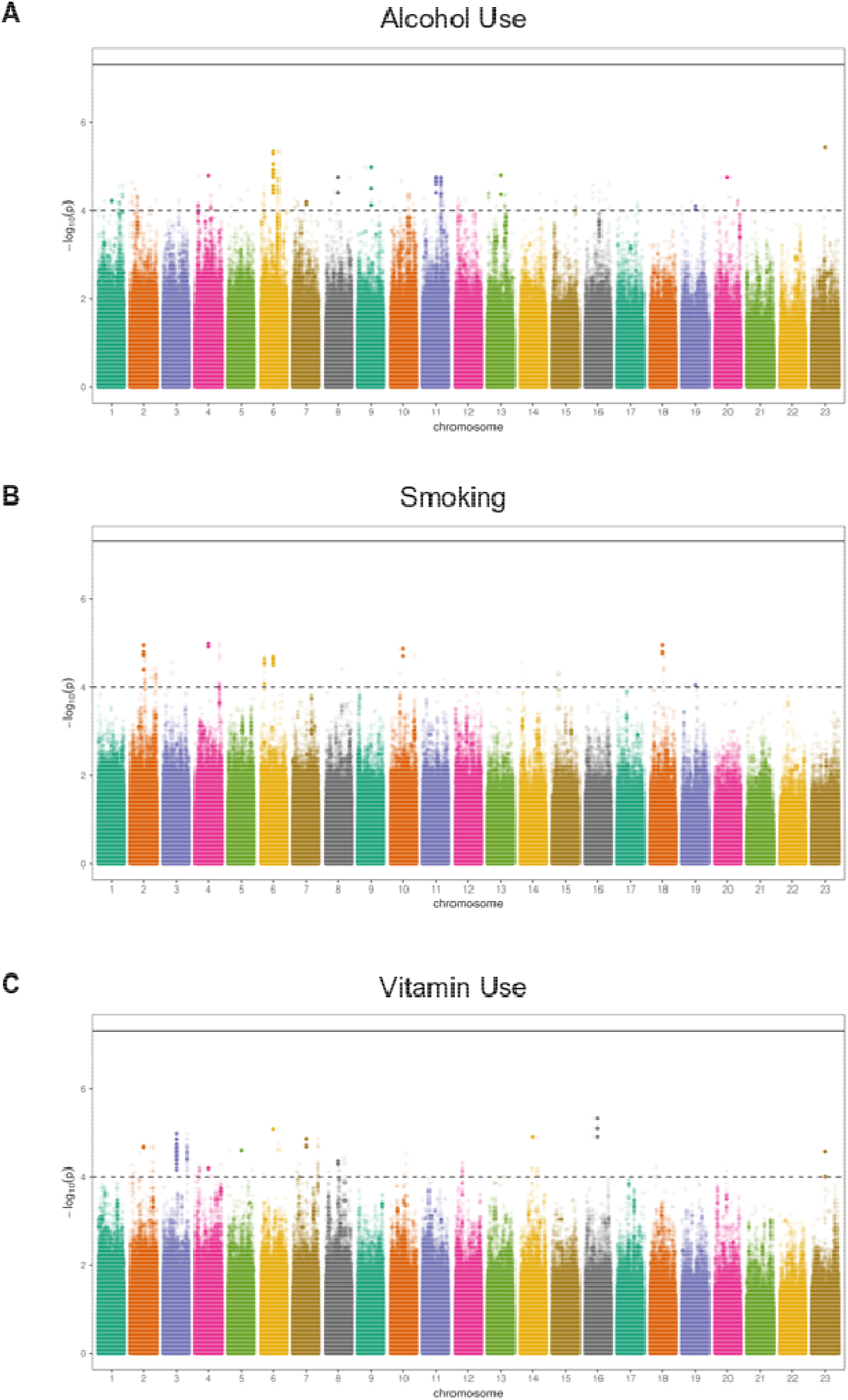
Manhattan plots for maternal periconceptional (A) alcohol use, (B) smoking, (C) vitamin use.

**Figure 2.**
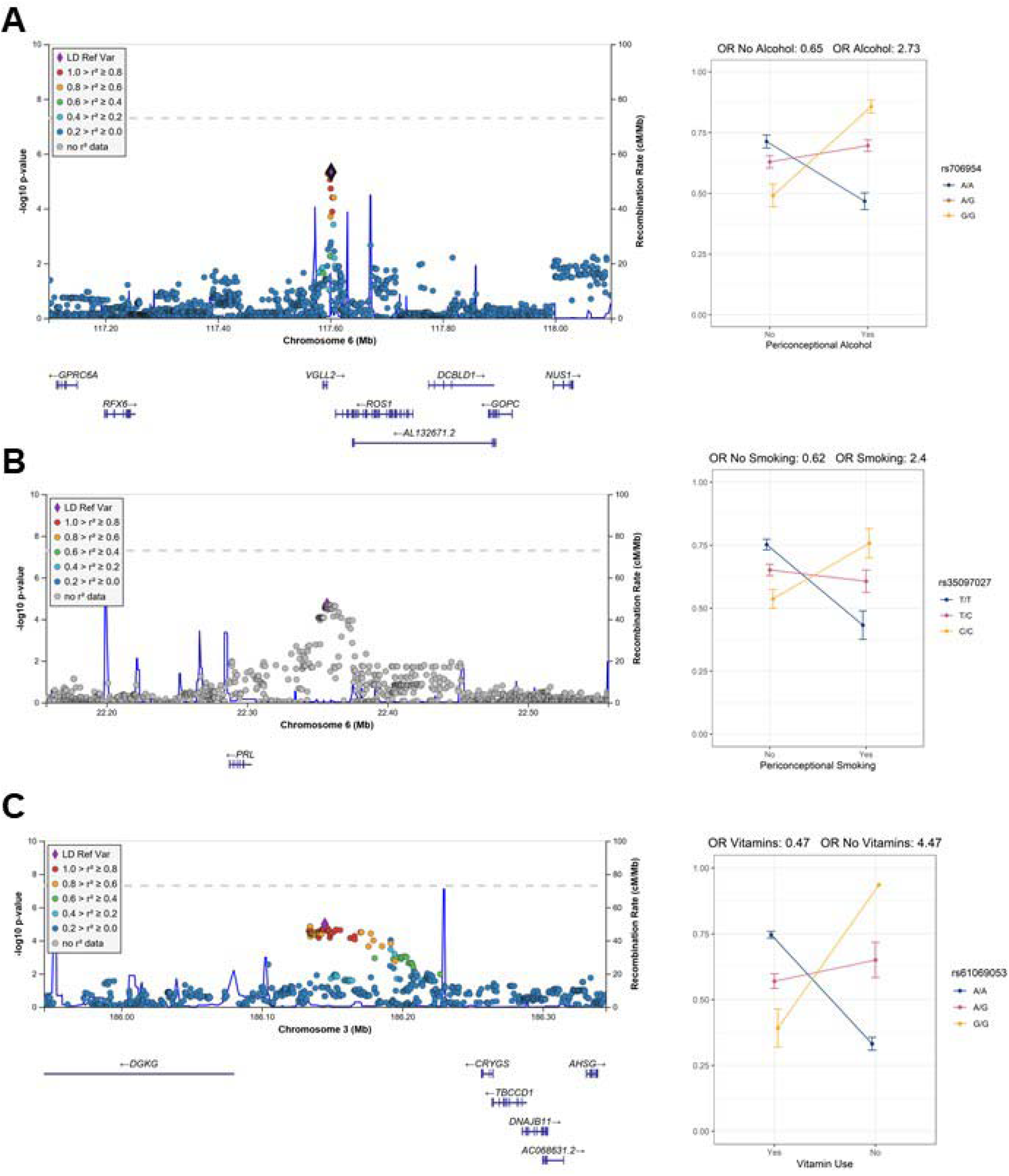
Regional association plots of the GE test with interaction plots. (A) 6q22 (rs706954) interaction with alcohol use. (B) 6p22.3 (rs35097027) interaction with smoking. (C) 3q27.3 (rs61069053) with vitamin use. Note, only one individual is homozygous for the G allele in the no vitamin use strata

Two additional loci had more than 5 SNPs per locus with significant GE p-values for alcohol exposures. These included 6q21 (lead SNP rs6929494, p_GE_=1.18×10^−5^) and 11q21 (lead SNP rs7116990, p_GE_=1.80×10^−5^). At the 6q21 locus, which included genes *REVL3* and *TRAF3IP2*, the major allele (A) of rs6929494 was associated with higher odds of NSCL/P within individuals with periconceptional alcohol use, but had an opposite (protective) effect for individuals without periconceptional alcohol use (Figure S1). The associated SNPs at the 11q21 locus, are located in an intergenic region between *CNTN5* and *JRKL*; for the lead SNP, rs7116990, the major allele (A) was associated with greater odds of NSCL/P in the individuals with periconceptional alcohol use and lower odds of NSCL/P for individuals without periconceptional alcohol use (Figure S1).

In the periconceptional smoking and vitamin use analyses, only one locus in each study had more than 5 significant SNPs. Thirty-six SNPs on 3q27.3 showed a significant interaction with vitamin use (lead SNP rs61069053, p_GE_=1.05×10^−5^). The minor allele (G) of rs61069053, located in an intergenic region between *CRYGS* and *DGKG*, had higher estimated odds of NSCL/P in the individuals without periconceptional vitamin use, but had lower odds of NSCL/P for the periconceptional vitamin user group (Figure S1). However, this result may be confounded by the distribution of vitamin use and allele frequency; only one individual was homozygous for the minor allele and did not have vitamin use. At 6p22.3, 47 SNPs showed significant GE p-values for smoking (lead SNP rs35097027, p_GE_=1.96×10^−5^). These variants are upstream of *PRL*; the minor allele (C) of rs35097027 conferred higher odds of CL/P for individuals with maternal smoking, but lower odds of NSCL/P for individuals without maternal smoking (Figure S1).

## DISCUSSION

The primary goal of this paper was to identify additional gene-environment interactions for NSCL/P and build on our previous work in this dataset (18). These analyses were motivated by multiple studies reporting associations between OFCs and periconceptional smoking, alcohol use, and vitamin supplementation (10, 23). Genome-wide interaction scans, however, have been limited for NSCL/P (13, 16). Our analyses add to a growing list of loci for which interactions between SNPs and maternal environmental exposures influence risk of OFCs.

The 2df joint G-GE test has statistical power above 80% for variants with minor allele frequency > 0.25 with strong environmental and/or genetic effects (OR>1.5), even with a relatively small sample size (21). Consistent with these calculations, the loci in these analyses with statistically significant G-GE associations that were driven by the GE effect had markedly strong effect estimates. Notably, the estimated odds of NSCL/P were 2.73 times higher with each copy of the minor allele G of rs117083 for individuals with periconceptional alcohol use and was 35% lower for individuals without periconceptional alcohol use. For the 5 loci with more than 5 SNPs per locus with significant GE p-values (i.e., 6q21 [alcohol], 6q22.1 [alcohol], 11q21 [alcohol], 6p22.3, [smoking], and 3q27.3 [vitamin use]), the genotypic odds ratio ranged from 60% lower odds of CL/P to 4.47 times higher odds. For alcohol and smoking exposures, we primarily identified scenarios described as “cross-over” interactions where the genotype group at lowest risk for NSCL/P without the exposure had high risk for NSCL/P with the exposure. In each of these interactions, the influence of genotype entirely depends on the exposure. The method we used to detect interactions is better suited for these “cross-over” interactions. However, it is limited in its ability to detect associations that are only present in one environmental context. For example, it is quite plausible that a genetic variant may only modify risk in the presence of a specific environment. This method has lower statistical power to detect such an association; additional methods are needed to explicitly search for these association signals.

We identified multiple loci with significant interactions. Of these, two have notable biological plausibility which we discuss in detail. One of the loci interacting with periconceptional alcohol use was on 6q22.1, approximately 5kb upstream of *VGLL2*, encoding vestigial-like 2. *VGLL2* is one of several Vestigial-like factors that plays a role in muscle fiber differentiation and the distribution of skeletal muscle fibers (24). Although its expression in adult tissues is restricted to skeletal muscle, during development, Vgll2 is more broadly expressed. In mice, its expression is enriched in the mandibular and maxillary prominences at embryonic day 10.5 and is localized to the epithelia of these structures(25). In the zebrafish, Vgll2 is expressed in the pharyngeal pouches and somites (26, 27). A role for *VGLL2* in craniofacial development is further supported by vgll2a zebrafish morphants, which induce death of neural crest cells in the pharyngeal arches causing hypoplasia of the Meckel’s and palatoquadrate cartilages and truncation of the ethmoid plate (25). Models for fetal alcohol syndrome indicate that ethanol exposure enhances cell death of cranial neural crest cells (28, 29). Although a specific interaction between VGLL2 and ethanol has not been described in vitro or in vivo, these data, combined with our statistical interaction suggest this may be a fruitful area for future studies.

At the 6p22.3 locus associated with maternal smoking, the lead SNP was located 60kb upsteam of *PRL*, which encodes prolactin. Prolactin is a reproductive hormone primarily known for its ability to stimulate mammary gland development and lactation, but it also has a variety of functions in reproduction, growth, and development (30). Prolactin receptor transcripts and proteins have been detected in non-lactogenic tissues notably including the facial cartilage and olfactory epithelium in mammals (31). Prolactin levels are known to be altered by both active smoking and exposure to secondhand tobacco smoke (32-35). Although smoking is associated with lower prolactin levels in pregnant women, the same is not true for fetal prolactin levels (36). A role for prolactin in craniofacial development and OFCs is unclear as knockouts of mouse PRL receptors are viable and lack gross morphological defects (37). However, in amphibian models, prolactin signaling has been suggested as the pathway underlying the ability of pre-metamorphic *Xenopus laevis* tadpoles to correct craniofacial defects induced by thioridazine (38). However, the same group also found that increased prolactin signaling during development alone does not cause craniofacial defects. Nonetheless, there is evidence linking smoking to prolactin levels and for prolactin signaling in craniofacial development; additional research will be needed to connect these results and determine a role for PRL and smoking in OFCs.

Our previous studies using this dataset have made use of the ancestral diversity that resulted from the 13 different recruitment sites worldwide. However, this study was limited to individuals of European descent as the number of unrelated participants with complete data from the other ancestry groups was quite small. In previous GWASs for gene-environment interactions, a lack of observations precluded the analysis of smoking and alcohol exposures in Asian trios (16).

The authors speculated that this was consistent with general trends of low alcohol and cigarette use among Asian women (39). This may also be true for other populations recruited into our study, but the incomplete data may also be due to changing study designs over the recruitment period. We also note that the recruitment of these participants was not population-based and thus does not reflect the population prevalence of NSCL/P nor the three environmental exposures we considered. In addition, a binary exposure variable does not capture or represent the range of possible environmental exposures that may influence development of NSCL/P. Therefore, the conclusions drawn here are limited to the study sample used in these analyses and cannot be used to draw conclusions at a broader level.

To summarize, we performed a genome-wide analysis to detect gene-environment interactions influencing risk of NSCL/P in Europeans. We identified two notable interactions: near VGLL2 and PRL with periconceptional alcohol use and smoking, respectively that are plausibly associated with risk of NSCL/P. These interaction effects are novel and warrant further investigations. If confirmed, these interactions provide new insights into the etiology of NSCL/P and could provide opportunities to modify risk through behavioral changes.

## Data Availability

All data are available from dbGaP

https://www.ncbi.nlm.nih.gov/projects/gap/cgi-bin/study.cgi?study_id=phs000774.v2.p1

## ACKNOWLEDGEMENTS

This project would not have been possible without the participation of families, field staff, and collaborators around the world. Special recognition to our colleague Dr. Andrew Czeizel (deceased). This work was supported by grants from the National Institutes of Health (NIH): R00-DE025060 [EJL], X01-HG007485 [MLM, EF], R01-DE016148 [MLM, SMW]. Genotyping and data cleaning were provided via an NIH contract to the Johns Hopkins Center for Inherited Disease Research: HHSN268201200008I.

## CONFLICT OF INTEREST

None.

## Notes

### Competing Interest Statement

The authors have declared no competing interest.

### Author Declarations

This study was approved by the IRBs at Emory University and the University of Pittsburgh.

